# Correlation Between Enzymatic and Non-Enzymatic Genes in *Acinetobacter baumannii* Isolates

**DOI:** 10.1101/2024.10.27.24316230

**Authors:** Alya Amer Rahi, Huda H. Al-Hasnawy

**Author notes:** **Corresponding Alya Amer Rahi**.

## Abstract

**Background:** *Acinetobacter baumannii* is a multidrug-resistant bacterium responsible for severe infections, particularly in hospital settings. Its resistance is driven by enzymatic genes such as those encoding beta-lactamases and carbapenemases, which degrade antibiotics, and non-enzymatic genes that modify mechanisms like efflux pumps and membrane permeability, further enhancing its defence against treatments. Together, these factors allow *A. baumannii* to thrive in clinical environments, complicating infection management.

**Objective:** This study aimed to explore the relationships between beta-lactamases, carbapenemases, efflux pumps, and membrane permeability changes, to understand their collective contribution to *A. baumannii’s* multidrug resistance.

**Materials and Methods:** Among 300 clinical isolates from urine, blood, wounds, and burns, 25 (8.33%) were identified as *A. baumannii*. These included 8% from urine, 12% from blood, and 40% each from wound and burn swabs. all specimens were taken from patients who have different symptoms in hospital of Al-Hilla Teaching Hospital/ Babylon. The research was carried out through the period January and June 2024. Bacterial identification was conducted using the VITEK-2 system and HI-Chromoagar® *A. baumannii*. Enzymatic genes were detected using conventional PCR, while non-enzymatic genes were analyzed via RT-qPCR.

**Results:** Molecular analysis revealed the presence of beta-lactamase (blaOXA-51, blaOXA-23) and metallo-beta-lactamase genes (blaVIM, blaIMP), with high antibiotic resistance rates. Gene expression analysis highlighted efflux pump upregulation (adeB) and altered permeability (CarO), reinforcing multidrug resistance mechanisms.

**Conclusion:** The combined action of enzymatic and non-enzymatic resistance genes in *A. baumannii* presents a significant treatment challenge, necessitating multi-target therapeutic approaches.

## Introduction

*Acinetobacter baumannii* is an opportunistic pathogen notorious for its role in nosocomial infections and its ability to rapidly develop resistance to multiple classes of antibiotics[^1^]. This gram-negative bacterium is associated with a variety of serious infections, including pneumonia, bloodstream infections, and wound infections, particularly in immunocompromised patients or those with prolonged hospital stays[^2^]. The alarming rise in multidrug-resistant (MDR) and extensively drug-resistant (XDR) strains of *A. baumannii* has made it a significant concern in healthcare settings worldwide[^3^]. Understanding the genetic mechanisms underlying its resistance is crucial for developing effective treatment strategies.Enzymatic genes in *A. baumannii* encode proteins that directly interact with antibiotics, often through enzymatic modification or degradation. The primary enzymatic mechanisms of resistance involve(Beta-Lactamases: These enzymes hydrolyze the beta-lactam ring of antibiotics such as penicillins and cephalosporins, rendering them ineffective[^4^]. *Acinetobacter baumannii* produces a range of beta-lactamases, including(Class A Beta-Lactamases: Such as TEM and SHV types, which primarily target penicillins and cephalosporins.Class B Beta-Lactamases: Including metallo-beta-lactamases (MBLs) like IMP and VIM, which can hydrolyze a broad spectrum of beta-lactam antibiotics, including carbapenems.Class D Beta-Lactamases: OXA-type beta-lactamases, which are particularly effective against carbapenems, contributing to the high levels of resistance seen in *Acinetobacter baumannii.*Carbapenemases A subset of beta-lactamases, carbapenemases, such as KPC and NDM types, are especially concerning due to their ability to hydrolyze carbapenems, often considered last-resort antibiotics[^5^]. The presence of carbapenemases in *Acinetobacter baumannii* isolates is a key indicator of extreme drug resistance.Non-enzymatic genes contribute to antibiotic resistance through mechanisms that do not involve direct enzymatic degradation of drugs. Non-enzymatic mechanisms include(Efflux Pumps: These membrane proteins actively transport antibiotics out of the bacterial cell, reducing their intracellular concentration and effectiveness[^6^]. Major efflux pump systems in *Acinetobacter baumannii* include(AdeABC: A well-studied efflux pump system in *Acinetobacter baumannii* that contributes significantly to resistance against multiple antibiotics, including carbapenems.Porins: These are membrane proteins that form channels through which antibiotics must pass to reach their targets inside the cell. Mutations or deletions in porin genes can decrease membrane permeability, reducing the uptake of antibiotics[^7^]. For instance, changes in the expression of the OmpA and CarO porins have been associated with reduced susceptibility to carbapenems[^8^].

## Material and Methods

### Clinical Specimens & Cultural Characteristics

A cross-sectional study was conducted involving 300 clinical specimens collected from patients who attended to hospitals in Hilla city: Al-Hilla General Teaching from January 2024 to June 2024. They’ve been collected from wounds, burns, urine, blood.When the samples was collected, it was immediately transported to the laboratory. Streaking of samples was done in MacConkey agar as well as nutrient agar, which were then incubated at 37°C in the aerobic environment for 24 hours. Bacterial colonies based on different morphology were individually isolated and treated with gram staining to be examined under a light microscope. These suspect colonies were later subcultured on HI-Chromagar agar also for 24 hours at 37°C.The bacteria growth and colorations were documented after using the Vitech 2 compact system. This approach was multifaceted that entailed the assessment and classification of *Acinetobacter baumannii* observed in the specimens taken from the patients.

### VITEK 2 system

The VITEK 2 Compact Instruments is an identification system, and this is a phenotypic type of identification that depends on biochemical reactions to identify the isolates. All isolates were inoculated onto MacConkey agar plates and then incubated overnight at 37°C. A single colony was used for identification by the VITEK-2 Systems method, done according to the manufacturer’s instructions.

### Antibiotic Susceptibility Test

All the 25 *A. baumannii* isolates were tested for antibiotic susceptibility, using the automated VITEK 2 Compact system. All samples were cultured on MacCkonkey agar plates, and then, a McFarland 0.5 standard suspension in 0.45% sodium chloride was prepared for every isolate. Liquid suspension of all isolates were loaded on the VITEK system and left overnight to get the result. 14 types of antibiotic were depended as the following included within the VITEK 2 Compact system Gram Negative Susceptibility card.

### DNA Isolation and Amplification

The genomic DNA of Isolated bacteria was extracted using the classical protocol by Presto Mini gDNA Bacteria Kit.

### Primers

The primers utilized in this study were produced by Macrogen company located in Korea. To prepare the working solution, the primers were diluted from stock using TE buffer to achieve a concentration of 10 picomoles per microliter, after which they were stored at -20°C. The oligonucleotide primers for all genes investigated in this research were sourced from prior studies and are detailed in Table (1). This table includes the primer sequences for each studied gene, along with their corresponding amplicon size in base pairs (bp) and the respective reference. These PCR primers were employed for detecting subtypes of system in clinical isolates of *A. baumannii*.

**Table (1):**
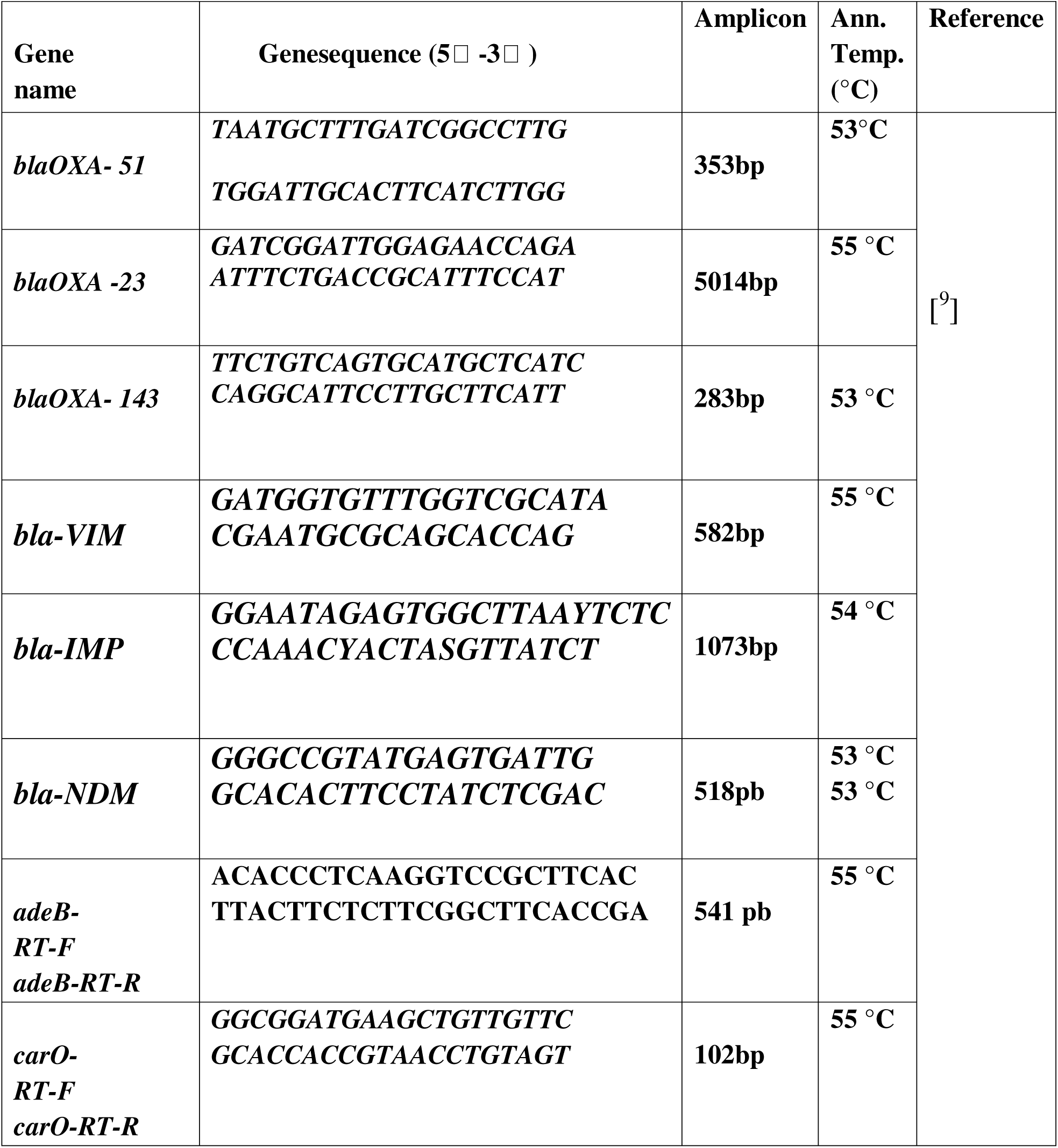
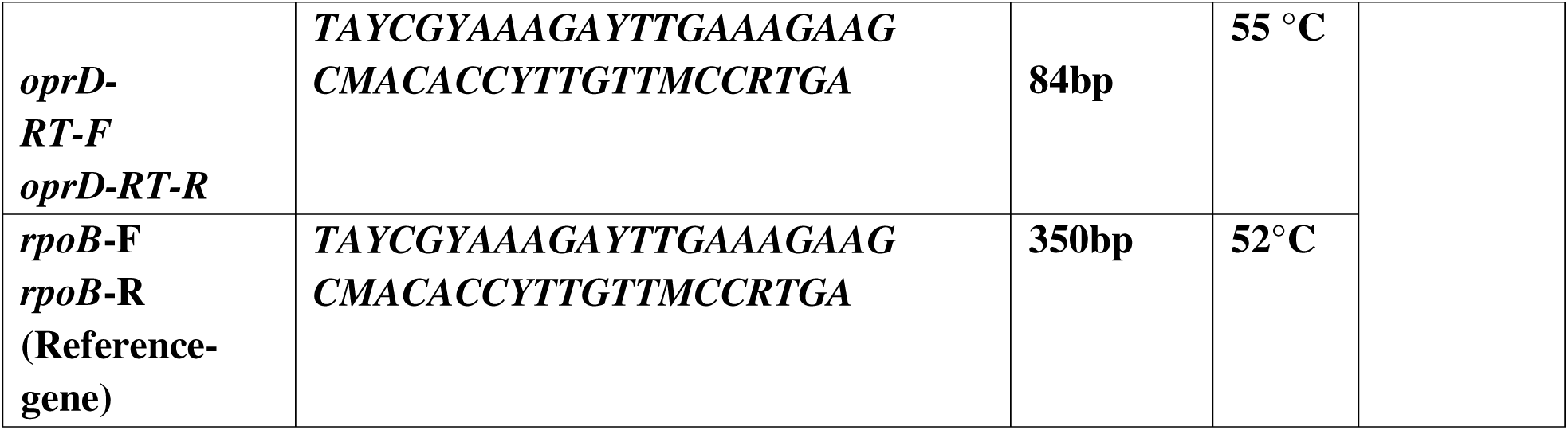
Primer sequences and their amplicon, annealing temperature in this study.

### Molecular detection of Enzymatic genes

The bacterial DNA was extracted by the alkaline lysis method. The pairs of primers (OXA-51, OXA-23, OXA-143,VIM,NDM,IMP) were used for PCR amplification genes. The amplification procedure was performed using 25 μl of master mix containing 0.2 μl of Taq polymerase 5 U/μl, 2.5 μl of 10XPCR buffer along with MgCl2, 1 μl of 10 pM from each reverse and forward primers, 2.5 μl of dNTPs MIX (2 Mm), 3 μl of DNA template, 14.8 μl of DNase-Free and RNase-Free Distilled Water. PCR amplification was performed using a thermal cycler. The amplified DNA products were then subjected to agarose gel electrophoresis, using a 1% agarose gel with a 100 bp size marker, for 10 minutes at 70 V. The gel was stained with ethidium bromide to visualize and detect the bands. The PCR reactions included both positive and negative controls. Table (1) shows the primers and their sequences used in this study.

### Real-Time qRT PCR Technique for Detection Non-Enzymatic Genes

Real-time quantitative PCR evaluated the expression of the pump for salt (adeB, adeJ) and the porin (oprD, carO) genes in 25 different isolates using SYBR Green I. EachPCR reaction included primers, cDNA, a SYBR Green I mixture, and water that was free of nucleases, with a final volume of 10 µL. The PCR conditions included a initial temperature of 94 °C for 5 minutes, followed by 40 cycles of 94 °C for 20 seconds, 60 °C for 20 seconds, and 72 °C for 30 seconds. A melt analysis that confirmed the presence of a single amplicon. The reference strain was Acinetobacter baumannii from the ATCC collection, and the rpoB gene was used as the internal control. All reactions were repeated three times, and the average CT value of each was used to calculate the gene’s expression level.

### Extraction of total RNA

Bacterial strains were cultured on MHA plates at 37 °C for 12 hours, and total RNA was extracted from the cultures during the late log phase using the High Pure RNA Isolation Kit. To verify the authenticity of the samples, DNA contamination was checked by omitting reverse transcriptase (RTase) from the controls. cDNA was synthesized from the RNA using reverse transcriptase and random hexamer primers provided in the kit. The cDNA was stored at −20°C for subsequent use in RT-PCR.

### Detection of adeB, CarO, OprD genes by Quantitative Real-Time PCR

In this study, we determine the fold of expression for adeB, CarO, OprD genes by determining delta CT by decreasing the mean CT of each isolate from the mean CT of the housekeeping gene and the delta CT was determined by decreasing the delta CT of each isolate from the delta CT of the control. The fold of expression was determined from the equation 2-(ΔΔCt).

### Correlation Between Enzymatic genes and Non-Enzymatic Genes

*Acinetobacter baumannii* demonstrates a complex resistance profile influenced by gene expression, particularly the OperD, CarO, and adeB genes. OperD is upregulated in isolates carrying blaOXA-51, blaOXA-23, blaVIM, and blaIMP, indicating a correlation with β-lactamase genes. CarO is downregulated in these same isolates, reducing antibiotic permeability. The adeB gene, crucial for the AdeABC efflux pump, is predominantly upregulated, contributing to multidrug resistance.

### Statistical analysis

The (2-ΔCT) method was employed to calculate the relative expression of genes. The rpoB gene (housekeeping) was employed as the endogenous control and was relative to the A.baumannii ATCC 19606 strain that served as the reference strain for the gene’s expression level. An RQ value of 1 indicates that the tested gene has a similar expression level in the reference and test strains, and an increase or decrease of 2 or 0.5 is considered significant. After Log2 fold change distribution was performed. Descriptive statistics for fold change gene expression represented by the mean, standard error (SE), standard division (SD), median and first-interquartile range (IQR), and minimum and maximum values. Categories variables expressed as frequencies and percentages and analyzed with chi-square or Fisher Exact Test. The differences in means rank among three gene-fold change gene expression was assessed by Kruskal-Wallis H. All data using SPSS 28.0 statistical software (SPSS Inc. Chicago, IL. USA). In addition, made of graphics with Excel 2021. A statistically significant difference was considered as a P-value < 0.05.

### Ethical approval

Ethical approval for this study was obtained from the ethical committee at Hilla Surgical Teaching Hospital. Furthermore, all individuals participating in the study were informed about the research, and their consent for both conducting the experiments and publishing the results was obtained prior to sample collection. This study was also approved by a local ethics committee at the College of Medicine, University of Babylon. and hospital ethics committee under document number [ IRB: 4-27, 3/1/2024].

## Result

### Distribution of *A.baumannii* in Different Specimens

The distribution of ***Acinetobacter baumannii*** isolates is based on their clinical specimen sources. The data are analyzed using a Chi-square test, yielding a Chi-square value of **9.08** with a **p-value of 0.028**, indicating statistical significance (p < 0.05). The distribution of isolates is as follows:

- **Burn swab:** 40% (10 isolates)
- **Wound swab:** 40% (10 isolates)
- **Blood:** 12% (3 isolates)
- **Urine:** 8% (2 isolates)

**Figure(1):**
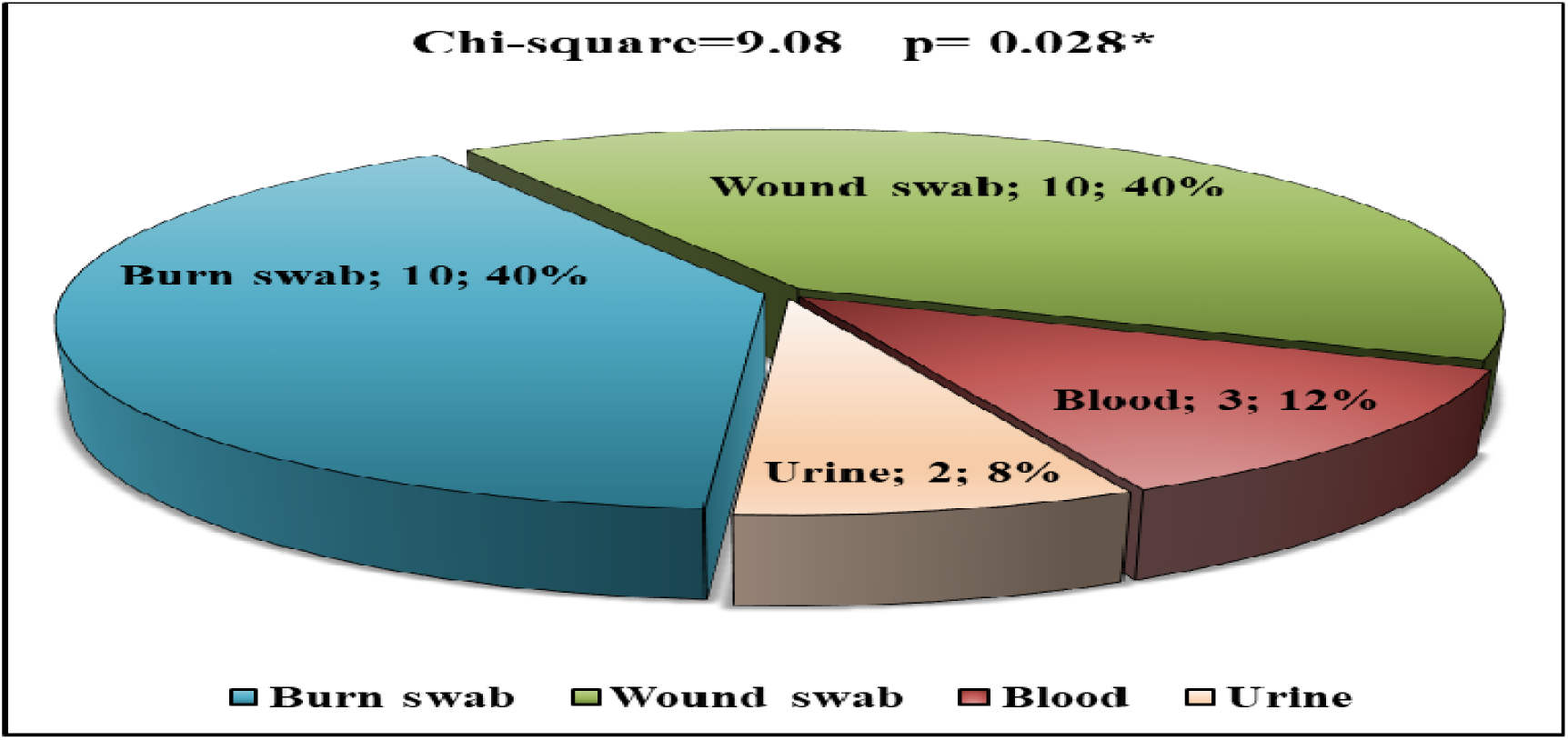
Distribution of *Acinetobacter baumannii* among Different Clinical Specimens.

### Vieck-2system

In this study, the biochemical tests done by Viteck -2 system, revealed positive reactions in key assays such as ADH1, BGLU, NC6.5, dMAN, and OPTO, highlighting its complex resistance mechanisms as in Table (2).

**Table (2).**
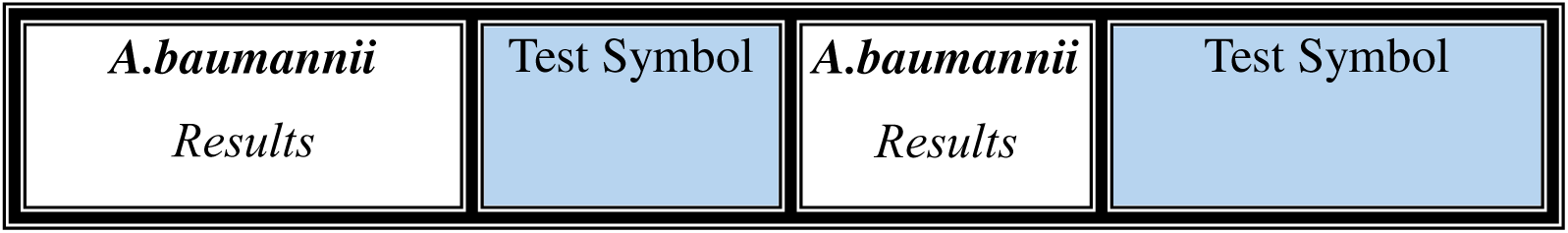
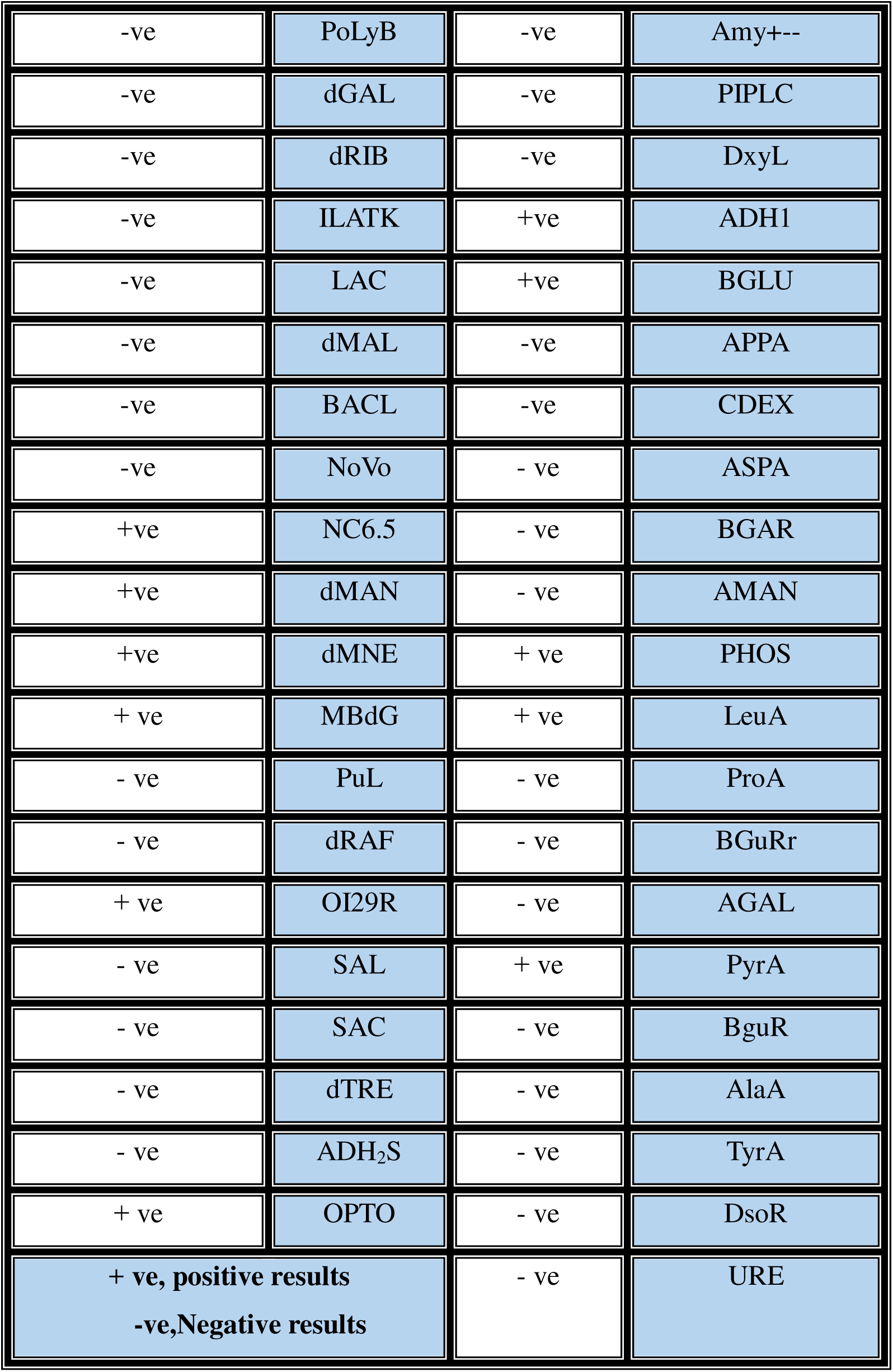
The Biochemical Tests by Viteck -2 system in 25 *A.baumannii*.

### Antimicrobial Susceptibility of 25 *A. baumannii* Isolates to Different Antibiotics

*Acinetobacter baumannii* exhibits significant antimicrobial resistance, as seen in 25 isolates tested in Table (3). High resistance rates were observed for Piperacillin/tazobactam, Colistin, Meropenem, and other antibiotics, emphasizing the challenges in treating infections caused by this pathogen.

**Table (3):**
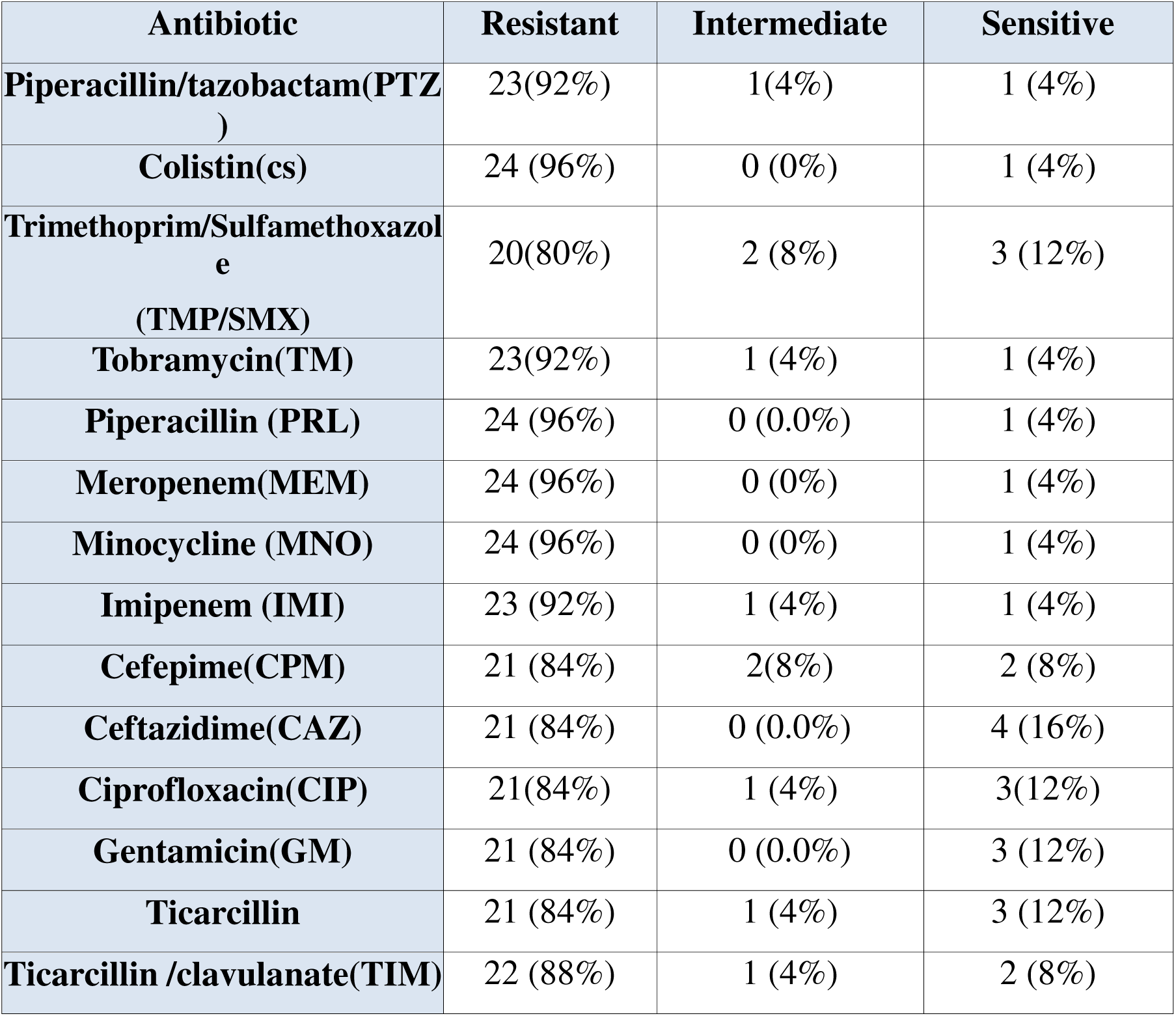
Antimicrobial Susceptibility Test of 25 *A. baumannii* Isolates.

### Molecular Detection of Enzymatic genes

The results of this study provide significant insights into the prevalence of oxacillinase and metallo-β-lactamase genes among *Acinetobacter baumannii* isolates recovered from clinical specimens as in Fig(2). The findings are consistent with global trends highlighting the rising concern of multidrug-resistant *A. baumannii* in healthcare settings.Oxacillinase Genes:The blaOXA-51 gene was detected in 100% of the isolates (25/25). This is consistent with the fact that blaOXA-51 is an intrinsic gene found in A. *baumannii*, serving as a marker for species identification. Its ubiquitous presence in all isolates supports the species’ identification and suggests that blaOXA-51 is a common feature of this bacterium, irrespective of resistance levels.The blaOXA-23 gene was identified in 80% of the isolates (20/25). This gene is associated with carbapenem resistance, which is one of the most significant challenges in treating infections caused by *A. baumannii.* The high prevalence of blaOXA-23 in this study indicates a considerable level of carbapenem resistance among the isolates, consistent with reports from other regions experiencing a surge in carbapenem-resistant *A. baumannii* (CRAB).Interestingly, none of the isolates carried the blaOXA-143 gene, which is another oxacillinase gene reported in some regions. The absence of blaOXA-143 suggests that this variant may not be prevalent in the studied population or that its distribution might be geographically restricted.Metallo-β-lactamase (MBL) Genes:The blaVIM gene, one of the MBL genes associated with resistance to a broad range of β-lactams, was present in 68% of the isolates (17/25). This suggests a substantial presence of MBL-producing *A. baumannii* in the clinical environment, complicating the treatment options as MBLs can hydrolyze carbapenems, one of the last-resort antibiotics.The blaIMP gene was detected in 60% of the isolates (15/25), further indicating the presence of MBL-producing isolates. The co-occurrence of blaVIM and blaIMP in a significant proportion of the isolates underscores the multidrug-resistant nature of these strains and highlights the need for stringent infection control measures.Notably, the blaNDM gene, another important MBL gene, was absent from all the isolates. The absence of blaNDM suggests that this gene may not yet be widespread in the studied population, or that other MBL genes, like blaVIM and blaIMP, are more predominant in this region.

**Figure(2):**
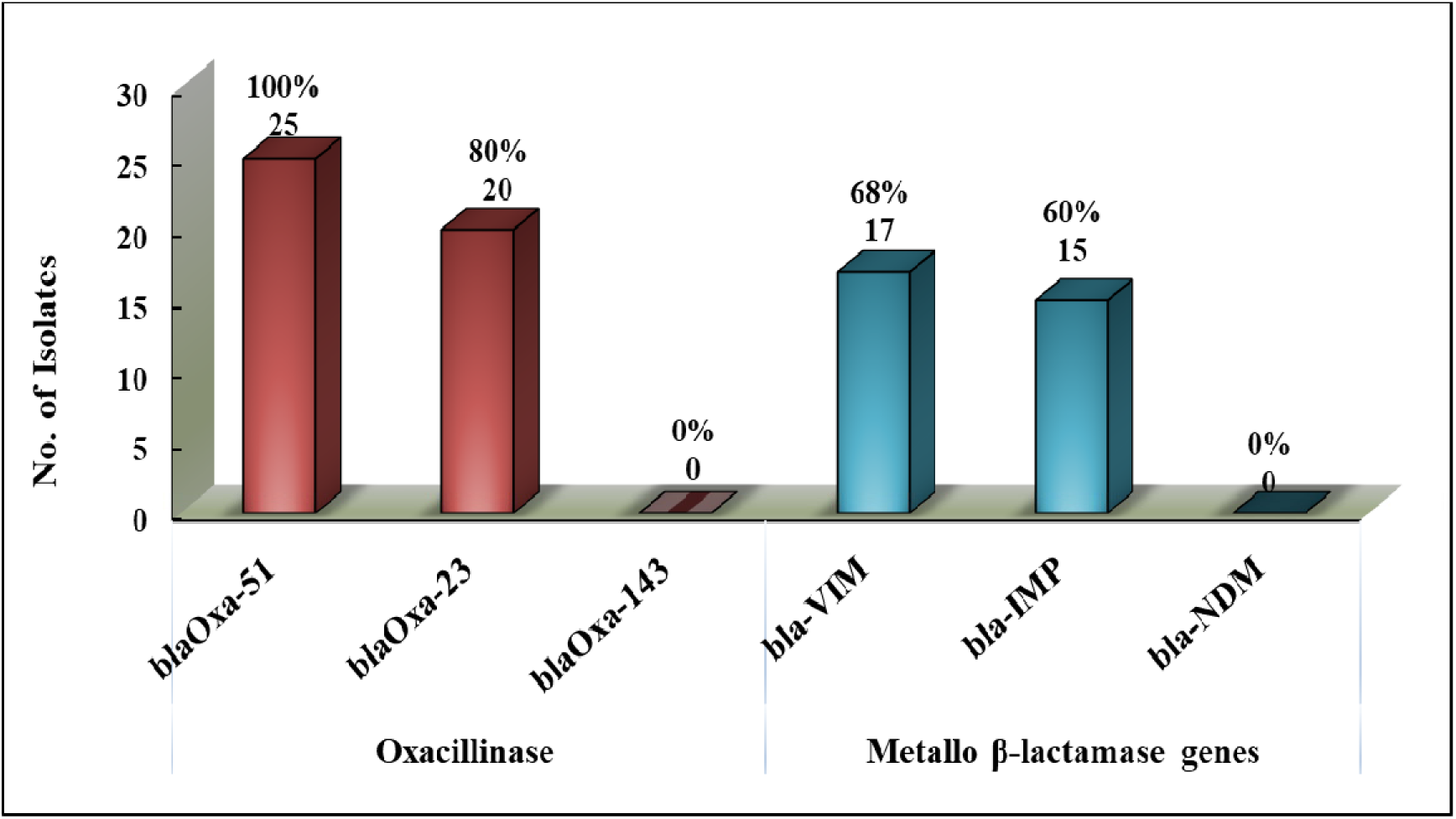
Percentage rate for Oxacillinase genes and Metallo B-Lactamase genes used in this study.

### Real-Time qRT PCR Technique for Detection of Non-Enzymatic Genes

In the present study the fold change expression of three genes (OperD, CarO, and adeB) in *Acinetobacter baumannii* isolates. The data provide insight into the differential expression of these genes, which are associated with antibiotic resistance mechanisms in *A. baumannii.* The expression of the OperD gene shows a significant fold change, with a mean value of approximately 10. This indicates that OperD is highly expressed in the studied isolates. The OperD gene has been implicated in resistance to multiple antibiotics, and its overexpression suggests that it may play a key role in the antibiotic resistance profiles of these isolates.The CarO gene exhibits extremely low expression, with a fold change near zero. CarO is a porin protein involved in the uptake of antibiotics, such as carbapenems, into the bacterial cell. The downregulation or loss of CarO expression could be a mechanism by which *A. baumannii* decreases its permeability to antibiotics, contributing to resistance, particularly to carbapenems. This finding is consistent with the behaviour of multidrug-resistant isolates that often exhibit porin loss to evade antibiotic action.The adeB gene also shows a notable level of expression, with a fold change of approximately 5. adeB is part of the AdeABC efflux pump system, which actively pumps out antibiotics from the bacterial cell, reducing their intracellular concentrations and effectiveness. The upregulation of adeB indicates that efflux pump-mediated resistance is another important resistance mechanism in these isolates.

**Figure(3):**
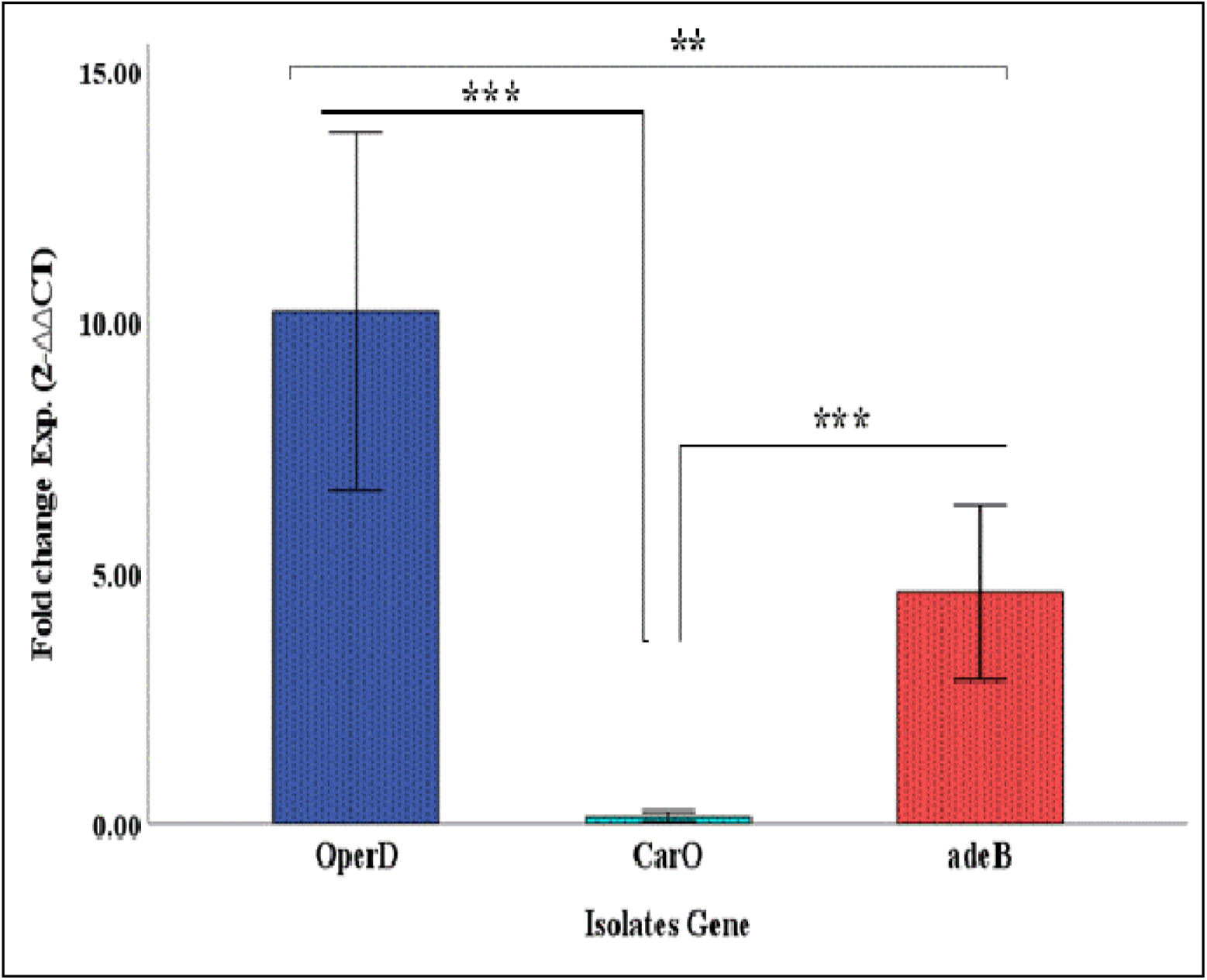
Fold change expression of three genes (OperD, CarO, and adeB) in *Acinetobacter baumannii* isolates.

### Correlation Between Enzymatic genes and Non-Enzymatic Genes

The correlation between gene expression and the resistance mechanisms through OperD Gene Expression:*blaOXA-51*, *blaOXA-23, blaVIM*, and *blaIMP* genes are associated with the upregulation of the OperD gene, indicating a strong correlation between the presence of these β-lactamase genes and the increased expression of *OperD.* Specifically:100% of isolates with *blaOXA-51* show upregulation of OperD.80% of isolates with blaOXA-23 show upregulation of *OperD*.68% of isolates with *blaVIM* and 60% of isolates with *blaIMP* also show increased OperD expression. The lack of *blaOXA-143* and *blaNDM* genes correlates with no significant effect on *OperD* expression. The absence of these genes in the isolates might suggest that OperD expression is more strongly linked to other β-lactamase genes in the clinical setting. The *CarO* gene shows complete downregulation in all isolates carrying *blaOXA-51*, *blaOXA-23, blaVIM, and blaIMP* genes. Specifically:100% of isolates with blaOXA-51 and *blaOXA-23* exhibit downregulation of CarO.68% of isolates with *blaVIM* and 60% of isolates with *blaIMP* also exhibit downregulation of CarO.This downregulation suggests that the presence of these β-lactamase genes, particularly *blaOXA-51* and *blaOXA-23*, is linked to reduced expression of the CarO gene. Since *CarO* encodes a porin involved in antibiotic uptake, its downregulation likely contributes to decreased permeability and increased resistance to antibiotics like carbapenems.No isolates showed upregulation of CarO, reinforcing its role in resistance through downregulation rather than active efflux or antibiotic degradation.adeB Gene Expression:The adeB gene is generally upregulated in isolates carrying *blaOXA-51, blaOXA-23, blaVIM,* and *blaIMP*. This suggests that the AdeABC efflux pump system, where adeB plays a central role, is active in conjunction with these β-lactamase genes, contributing to the multidrug-resistant phenotype.100% of isolates with *blaOXA-51* showed adeB upregulation.81.8% of isolates with *blaOXA-23* and 72.7% of isolates with *blaVIM* exhibited upregulation of *adeB.*Although a small percentage of isolates (3 isolates) showed downregulation of adeB, the overwhelming trend is towards upregulation in the presence of these β-lactamase genes.Similar to *OperD,* the adeB gene is not affected by *blaOXA-143* and *blaNDM*, which were absent in the isolates, suggesting that its expression is linked to the other β-lactamase genes.

**Table (4):**
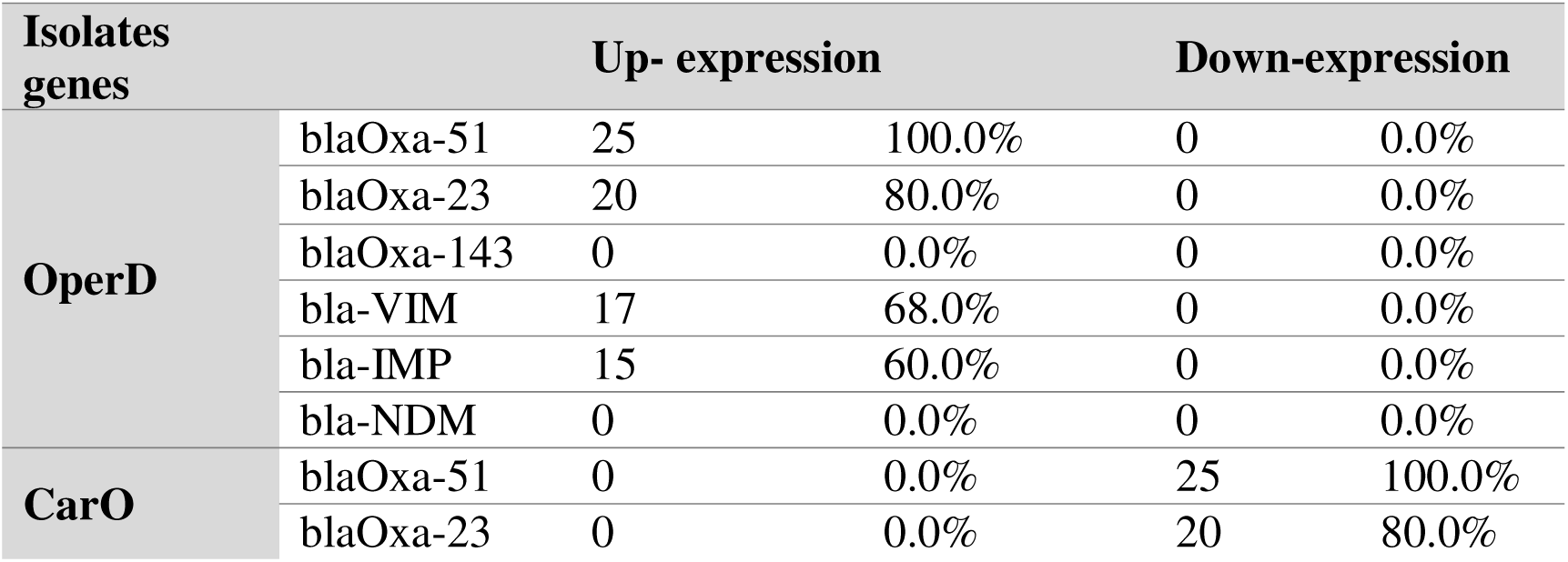
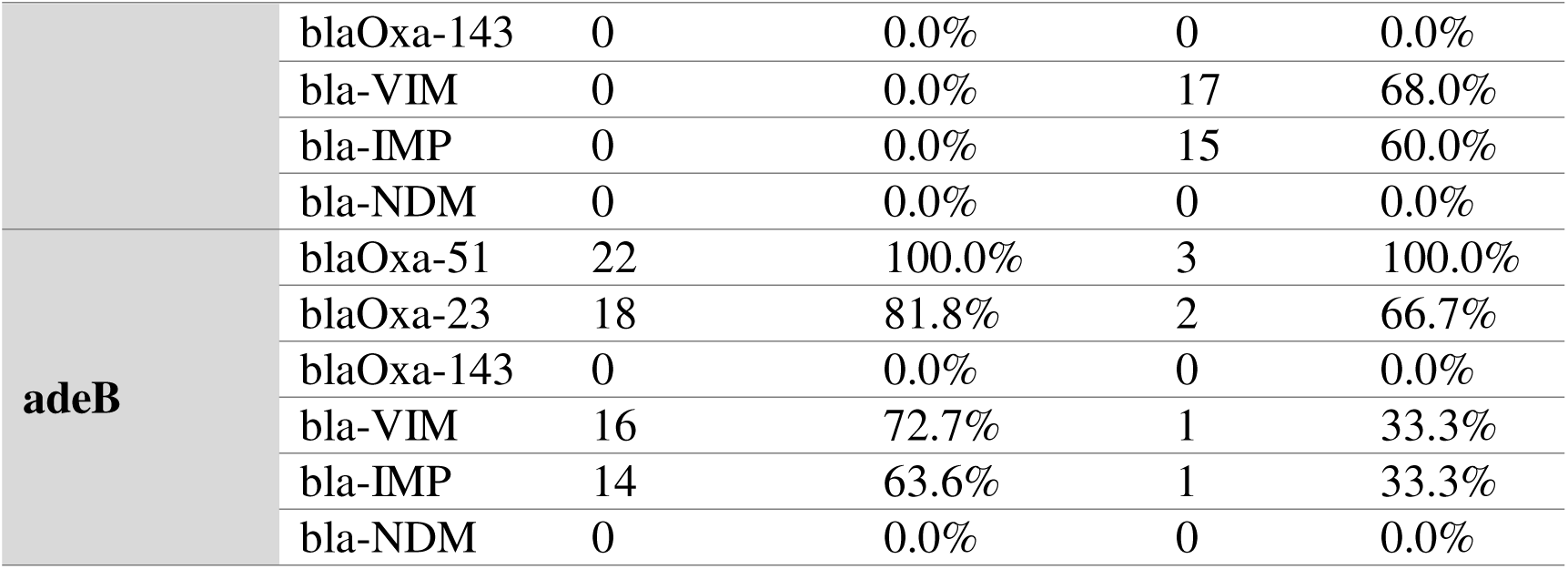
Correlation Between Enzymatic genes and Non-Enzymatic Genes.

## Discussion

The distribution of *Acinetobacter baumannii* isolates across different clinical specimens showed a statistically significant variation, as demonstrated by the Chi-square test (χ² = 9.08, p = 0.028). Burn swabs and wound swabs each contributed 40% of the isolates, while blood and urine samples accounted for 12% and 8% of the isolates, respectively. This distribution reflects the opportunistic nature of *A. baumannii*, which thrives in environments where skin and soft tissue infections are common, such as burn units and surgical sites. The lower percentages in blood and urine samples suggest that while A. *baumanni*i can cause systemic infections and urinary tract infections, its prevalence in these areas is less compared to wound and burn infections. This data emphasizes the importance of closely monitoring burn and wound patients, as they represent significant reservoirs for *A. baumannii* in healthcare settings. Agree with the study [^10^] found 250 *A. baumannii* isolates were distributed as 6(50%) recovered from urine, 4(33.3%) from lower respiratory secretion and only two (25%) were obtained from wound pus, no *A. baumannii* isolates were isolated from blood samples in this study,and study by [^11^]a total of 664 samples were collected from patients in different Specimens. The specimens analyzed included blood (122), catheter (34), pus (198) and pulmonary (310). The antimicrobial susceptibility testing revealed alarmingly high resistance rates among the 25 *A. baumannii* isolates. Notably, 96% of the isolates were resistant to Colistin, a last-resort antibiotic for multidrug-resistant (MDR) Gram-negative infections. Resistance to Piperacillin/tazobactam (92%), Meropenem (96%), and Imipenem (92%) further underscores the challenges posed by MDR *A. baumannii* in clinical settings. The high resistance rates to carbapenems, coupled with substantial resistance to aminoglycosides (e.g., Tobramycin, Gentamicin), fluoroquinolones (e.g., Ciprofloxacin), and β-lactam/β-lactamase inhibitor combinations (e.g., Piperacillin/tazobactam, Ticarcillin/clavulanate), reflect the limited treatment options available for infections caused by this pathogen. This necessitates a more stringent antibiotic stewardship approach and the development of alternative therapeutic strategies. Agree with study by [^12^] who founded the results showed that phenotypic resistance against amikacin was 27.2%, ceftriaxone 100%, ceftazidime 27.2%, cefepime 63.3%, ciprofloxacin and co-trimoxazole 100%, gentamicin 40%, imipenem 22.2%, meropenem 21.1%, piperacillin-tazobactam 27.2%, tigecycline 27.2%, and tetracycline 63.3%. All *A. baumannii* isolates were found to be sensitive to colistin (CT), polymixin-B (PB), and tobramycin (TOB).The study by [^13^] found that 25 *A.baumannii* isolates against fourteen types of antibiotics were equal to cefotaxime (100 %), ticarcillin-clavulanate, and ceftriaxone (96 %), and (92 %) for each of Cefepime and Tobramycin, the lowest percentage was for doxycycline (64 %). The molecular analysis revealed that the *blaOXA-51* gene, an intrinsic oxacillinase marker for *A*. *baumannii*, was present in 100% of the isolates. *The blaOXA-23* gene, associated with carbapenem resistance, was detected in 80% of the isolates, indicating a significant burden of carbapenem-resistant A. baumannii (CRAB) in the studied population. The absence of *blaOXA-143* and *blaNDM*\* genes suggests that these resistance determinants are either geographically restricted or less prevalent in this specimens, as opposed to *blaOXA-51* and *blaOXA-23*, which appear to be dominant. The presence of metallo-β-lactamase (MBL) genes, including blaVIM (68%) and blaIMP (60%), further complicates treatment, as MBLs confer resistance to a broad range of β-lactam antibiotics, including carbapenems. The absence of the blaNDM gene, another significant MBL gene, may indicate that this resistance mechanism is not yet widespread in the specimens. Agree with the study [^14^] found In Acinetobacter baumannii Thirty-five (80%) of Forty-four presumptive MBL producer isolates were positive for bla IMP-1, while Twelve (27.3%) Acinetobacter baumannii of 44 presumptive MBL producer isolates were positive for blaIMP gene and all isolates carrying *blaOxa-51* and 80% carrying blaOxa23 and study by [^15^] who founded 172 isolates carrying *blaOXA-51* and *blaOXA-23* genes were detected in all isolates, while blaOXA-134, were not detected in all isolates. Further analysis showed that 31.97% of isolates co-harboured *blaVIM, blaIMP* and *blaNDM* in percentage 80%, and 72%.4% respectively. While The *OperD* gene showed significant upregulation, with a mean fold change of approximately 10, highlighting its potential role in resistance mechanisms, particularly in association with β-lactamase genes like *bla*OXA-51 and *bla*OXA-23. The upregulation of *OperD* may enhance the bacterium’s ability to survive in the presence of multiple antibiotics, making it a critical target for future therapeutic interventions.while study [^16^] found that OprD downregulation was observed in multidrug-resistant and pan-drug-resistant *A. baumannii*. Decreased membrane porin density (CarO) was associated with pan-drug resistance in *A. baumannii*. In present study The *CarO* gene, which encodes a porin involved in antibiotic uptake, exhibited downregulation in all isolates. This downregulation likely reduces the permeability of the bacterial cell membrane to antibiotics, particularly carbapenems, contributing to the high level of resistance observed in these isolates. The loss of *CarO* expression is a common mechanism in MDR *A. baumannii* and further complicates the treatment of infections caused by this pathogen.Agree with a study by [^17^] found all strains carrying *bla*_OXA-51_ like and *bla*_OXA-23_ like in 100%. Nonetheless, *bla*_OXA-134_ was harboured by 0 per cent in all strains. The relative expression levels of the *carO* gene ranged from 0.06 to 35.01 fold lower than that of carbapenem-susceptible *A. baumannii* ATCC19606 and analysis of the outer membrane protein showed that all 100 isolates produced CarO. The results of current study revealed the prevalence of *bla*_OXA_ genes and changes in *carO* gene expression in carbapenem-resistant *A.baumannii*.The *adeB* gene, a key component of the AdeABC efflux pump, also demonstrated notable upregulation in most isolates, indicating that efflux-mediated resistance is another significant mechanism in these isolates.And study by [^18^] found that all isolates *A.baumannii* have upregulated expression of the *adeB* agreeing with the present study. The correlation between the upregulation of *adeB* and the presence of β-lactamase genes further supports the multifaceted resistance mechanisms employed by *A. baumannii*.The correlation analysis revealed a strong association between the presence of β-lactamase genes (*bla*OXA-51, *bla*OXA-23, *bla*VIM, and *bla*IMP) and the upregulation of non-enzymatic resistance genes (*OperD* and *adeB*). This suggests that these genes work in concert to enhance the multidrug-resistant phenotype of *A. baumannii*. The downregulation of *CarO* in isolates carrying these β-lactamase genes further underscores the role of decreased permeability in resistance mechanisms. The absence of *bla*OXA-143[^19,20,21^] and *bla*NDM correlated with a lack of significant expression changes in *OperD*, *CarO*, and *adeB*, suggesting that these genes may not play a major role in the resistance profiles of the isolates[^22,23,24^].

## Conclusion

The findings of this study highlight the critical challenges posed by multidrug-resistant *A. baumannii* in clinical settings. The high prevalence of resistance genes, coupled with significant expression of non-enzymatic resistance mechanisms, underscores the need for improved infection control measures and the development of novel therapeutic strategies to combat this pathogen. The correlation between enzymatic and non-enzymatic resistance genes provides a deeper understanding of the complex resistance mechanisms in *A. baumannii*, paving the way for targeted interventions to mitigate the spread of MDR strains in healthcare environments.

## Data Availability

clinical isolates from urine, blood, wounds, and burns,

## Acknowledgements

Thanks to the staff of Al-Hilla Teaching Hospital and Dr.Ashwaq Al-Shuwailia.

## Authors’ contributions

The authors conceived and designed the experiment performed the experiment, and analyzed the data, participated in its design and coordination and helped to draft the manuscript. All authors read and approved the final manuscript.

